# The *CADM2* gene and behavior: A phenome-wide scan in UK-Biobank

**DOI:** 10.1101/2021.04.16.21255141

**Authors:** Joëlle A. Pasman, Zeli Chen, Jacqueline M. Vink, Michel C. Van Den Oever, Tommy Pattij, Taco J. De Vries, Abdel Abdellaoui, Karin J.H. Verweij

## Abstract

This phenome-wide association study examined SNP and gene–based associations of the *CADM2* gene with 242 psycho-behavioral traits (N=12,211–453,349). We found significant associations with 51 traits, many more than for other genes. We replicated previously reported associations with substance use, risk-taking, and health behavior, and uncovered novel associations with sleep and dietary traits. Accordingly, *CADM2* is involved in many psycho-behavioral traits, suggesting a common denominator in their biology.

---

In the last 15 years, genome-wide association studies (GWASs) have identified tens of thousands of associations between genetic variants and a range of human behavioral and physical traits. One gene that has popped up surprisingly often in behavioral GWASs is the Cell Adhesion Molecule 2 gene *(CADM2)*. Common variations (single nucleotide polymorphisms, SNPs) in the *CADM2* gene have been implicated in various traits, including personality^1^, cognition and educational attainment^2,3^, risk-taking behavior^4^, reproductive success^5^, autism spectrum disorders^6^, substance use^7,8^, physical activity^9^, and BMI/obesity^10^.

*CADM2* encodes a member of the synaptic cell adhesion molecules (SynCAMs) involved in synaptic organization and signaling, suggesting that alterations in *CADM2* expression affect neuronal connectivity. *CADM2* is abundant in brain regions important for reward processing and addiction, including the frontal anterior cingulate cortex^3^, substantia nigra, and insula^11^. Given its common appearance in GWASs and its central role in brain functioning, *CADM2* is a gene that warrants further exploration.

In this study we perform a phenome-wide association analysis (PheWAS), in which we test for associations of *CADM2* (on SNP and gene level) with a comprehensive selection of psycho-behavioral phenotypes as measured in the UK Biobank cohort. Results will provide insights about whether the role of *CADM2* is confined to a specific set of traits or is involved in a wider range of phenotypes. This will inform future studies on the function of *CADM2* and the neurobiological underpinnings of different psycho-behavioral traits. An additional advantage is that the multiple testing burden is reduced as compared to genome-wide studies, resulting in higher power levels.

UK Biobank is a nationwide study in the United Kingdom containing phenotypic and genetic information for up to 500,000 individuals^12^. We analyzed 12,211 to 453,349 UK Biobank participants with European ancestry for whom genetic and phenotypic data were available. About half (54.3%) of the sample was female, and mean age was M=56.8 (range 39-73, SD=8.0). We extracted the *CADM2* region 250 kb up- and downstream (bp 84,758,133 to 86,373,579 on 3p12.1, GRCh37/hg19) and selected 4,265 SNPs with call rate>95%, minor allele frequency >1%, and p-value for violation of Hardy-Weinberg equilibrium of *p*_*HWE*_>10^−6^ (QC details are described in Abdellaoui, 2020^13^).

We selected 242 psychological and behavioral phenotypes from the UK Biobank with an effective sample size above 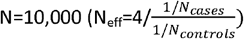. In order to maximize sample size, we used the first available measurement for each individual; if the first instance was not available, we took the second, otherwise the third, etc. In addition, we included eight traits that were derived for recent genetic studies, including seven substance use traits and educational attainment in years (for an overview of all included traits, see Supplemental Table S1). Continuous phenotypes were cleaned such that theoretical implausible values were set on missing and extreme values more than 4 SDs away from the mean were winsorized at the maximum value of 4SDs. Binary and ordinal variables were left unchanged. Ordinal variables were analyzed as continuous.

The SNP-based association analyses were performed in fastGWA^14^, taking into account genetic relatedness. Analyses were controlled for effects of age, sex, and 25 genetic principal components (PCs, to control for genetic ancestry^15^). We used linear mixed modeling for all traits and Haseman-Elston regression to estimate the genetic variance component. To test the significance of *CADM2*-associations on gene-level we conducted a MAGMA gene-based test^16^, which aggregates the SNP effects (regardless of direction) in a single test of association. We used the default SNP-wise mean procedure (averaging SNP effects across the gene) and checked the results of the SNP-wise top procedure for comparison (more sensitive when only a small proportion of SNPs has an effect). As significance threshold for the SNP-based test we adopted a genome-wide significance threshold of *p*<5E-08. As this is rather stringent given that we test within a single gene, we also used a significance threshold of .05 corrected for the number of independent SNPs (n=133, at R^2^=0.10 and 250kb) and the number of traits, resulting in .05/(133*242)=1.55E-06. For the gene-based test we used a threshold of 2.62E-05, corresponding to .05 divided by the total number of genes (19,082). To provide an estimation of the effect size, we used 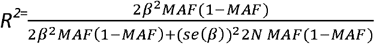, as described in ^17^, with adaptations for binary traits as described in^8^.

Given that our analyses were highly powered, we assessed whether the high number of associations discovered for *CADM2* was unusual or similar to those found for other genes. We therefore selected a random set of 50 genes (that were up to 50% smaller or larger in size), repeated the SNP-based analysis for these genes and compared the number of traits with significant associations..

On the SNP-level, 38 traits reached significant associations at a genome-wide corrected p-value (5E-08), and 61 traits at the lenient threshold of *p*<1.55E-06 (Figure 1a, Table 1). In the gene-based test, 51 out of 242 traits showed significant associations under a p-value of 2.62E-06 (Figure 1b, Table 1). The strongest associations were found for cognitive ability, risk taking, diet, BMI, daytime sleeping, sedentary behaviors, nervousness-like traits, reproductive traits, and substance use. There were fewer associations with occupational, traumatic experiences, social connection and non-worry related depression traits. Full SNP- and gene-based results are provided in Table S2 and S3a. Table S3b shows the gene-based results for the SNP-wise top procedure. There were some differences with the SNP-wise mean results, with only 33 significant associations and a correlation of r=.64 between the *p*-values from the respective tests.

**Table 1.** Phenotypes with a significant association with *CADM2* according to the MAGMA gene-based test (SNP-wise mean) at p<2.62E-06. The top-SNP for the phenotype is given with the minor allele (A1), beta (β), *p*-value (*p*), and percentage of explained variance in the respective trait (R^2^. (%)). Most top-SNPs were significant at *p*<1.55E-6 (bold-faced).

| Category | Variable label | N | $p(\text{Gene})$ | Top SNP | A1 | $\beta$ | $p$ | $R^2$ (%) |
| --- | --- | --- | --- | --- | --- | --- | --- | --- |
| Diet | Bread intake | 448,511 | 2.52E-15 | rs2326128 | A | 0.016 | **3.31E-14** | 0.013 |
| Diet | Fresh fruit intake | 451,787 | 7.27E-13 | rs12638798 | T | 0.015 | **1.42E-11** | 0.010 |
| Diet | Hot drink temperature | 448,701 | 3.61E-26 | rs17023019 | A | -0.012 | **5.21E-22** | 0.021 |
| Diet | Lamb/mutton intake | 450,807 | 3.98E-08 | rs10865611 | G | 0.007 | 3.24E-06 | 0.005 |
| Diet | Oily fish intake | 451,239 | 7.92E-14 | rs11712915 | C | 0.014 | **2.95E-10** | 0.009 |
| Diet | Salad / raw vegetable intake | 447,881 | 7.17E-13 | rs1248825 | A | -0.015 | **4.35E-11** | 0.010 |
| Diet | Salt added to food | 453,349 | 9.63E-25 | rs17516256 | G | 0.017 | **4.61E-20** | 0.018 |
| Exercise and sedentary behavior | Length of mobile phone use | 447,851 | 4.49E-08 | rs13092059 | A | 0.023 | 2.41E-06 | 0.005 |
| Exercise and sedentary behavior | Number of days/week of vigorous physical activity 10+ minutes | 431,716 | 5.98E-14 | rs2326123 | T | -0.030 | **3.96E-11** | 0.010 |
| Exercise and sedentary behavior | Time spend outdoors in summer | 428,647 | 3.09E-21 | rs62250754 | G | 0.019 | **1.27E-18** | 0.018 |
| Exercise and sedentary behavior | Time spent driving | 446,804 | 5.41E-07 | rs7609594 | G | 0.010 | 3.39E-06 | 0.005 |
| Exercise and sedentary behavior | Time spent outdoors in winter | 428,625 | 1.24E-16 | rs62252461 | A | -0.016 | **1.23E-13** | 0.013 |
| Exercise and sedentary behavior | Time spent using computer | 450,226 | 2.33E-07 | rs7642644 | C | -0.015 | **4.44E-07** | 0.006 |
| Exercise and sedentary behavior | Time spent watching television (TV) | 449,939 | 4.53E-14 | rs9824301 | C | -0.018 | **4.23E-16** | 0.015 |
| Exercise and sedentary behavior | Usual walking pace | 450,769 | 2.28E-06 | rs2290338 | T | -0.008 | **6.15E-07** | 0.006 |
| Mental health | Frequency of tiredness / lethargy in last 2 weeks | 440,102 | 8.70E-08 | rs818215 | C | -0.011 | **1.18E-10** | 0.010 |
| Mental health | Irritability | 433,487 | 1.24E-09 | rs6800177 | T | 0.007 | **3.33E-11** | 0.010 |
| Mental health | Nervous feelings | 441,742 | 3.59E-24 | rs1449386 | T | -0.008 | **7.93E-19** | 0.018 |
| Mental health | Neuroticism score | 367,273 | 8.97E-09 | rs818219 | C | -0.012 | **1.59E-07** | 0.008 |
| Mental health | Seen doctor (GP) for nerves, anxiety, tension or depression | 450,407 | 1.30E-06 | rs12631564 | A | 0.005 | 2.77E-06 | 0.005 |
| Mental health | Suffer from 'nerves' | 436,983 | 2.37E-19 | rs7652808 | T | -0.007 | **2.21E-14** | 0.013 |
| Mental health | Tense / 'highly strung' | 439,327 | 5.55E-12 | rs9811546 | A | -0.005 | **5.28E-08** | 0.006 |
| Mental health | Worrier / anxious feelings | 441,805 | 1.64E-17 | rs62250713 | A | -0.010 | **2.18E-20** | 0.019 |
| Physical health | Body mass index (BMI) | 452,241 | 2.22E-26 | rs11915747 | G | -0.019 | **1.40E-18** | 0.017 |
| Physical health | Childhood sunburn occasions | 339,817 | 2.07E-08 | rs9880919 | A | 0.017 | **3.00E-09** | 0.010 |
| Physical health | Mother's age at death | 275,917 | 4.20E-07 | rs9843797 | T | -0.014 | **1.07E-07** | 0.010 |
| Reproductive | Age first had sexual intercourse | 398,884 | 4.35E-27 | rs62263912 | G | -0.026 | **1.36E-29** | 0.033 |
| Reproductive | Lifetime number of sexual partners | 372,178 | 6.04E-13 | rs4856598 | A | 0.014 | **3.29E-09** | 0.010 |
| Reproductive | Number of children fathered | 205,843 | 4.75E-20 | rs1368750 | T | 0.029 | **1.14E-19** | 0.040 |
| Reproductive | Number of live births | 245,757 | 1.76E-09 | rs1972994 | A | 0.019 | **7.41E-11** | 0.017 |
| Risk taking | Risk taking | 437,513 | 2.12E-25 | rs6762267 | C | 0.011 | **1.27E-30** | 0.030 |
| Sleep | Daytime dozing / sleeping (narcolepsy) | 451,759 | 7.68E-11 | rs960986 | T | -0.007 | **2.86E-11** | 0.010 |
| Sleep | Nap during day | 453,179 | 2.30E-07 | rs3943782 | G | 0.007 | **5.89E-08** | 0.007 |
| Social interaction | Frequency of friend/family visits | 451,075 | 1.65E-06 | rs1248860 | G | 0.011 | **1.05E-06** | 0.005 |
| Socioeconomic status and intelligence | Average total household income before tax | 390,512 | 2.37E-07 | rs426444 | T | -0.015 | **4.59E-08** | 0.008 |
| Socioeconomic status and intelligence | Educational attainment transformed to ISCED categories, derived for PMID 27225129 | 449,507 | 1.21E-19 | rs11915747 | G | 0.095 | **3.86E-18** | 0.017 |
| Socioeconomic status and intelligence | Fluid intelligence score | 233,219 | 3.25E-08 | rs72903244 | A | -0.047 | **2.91E-08** | 0.013 |
| Socioeconomic status and intelligence | Number in household | 450,782 | 3.52E-07 | rs62250661 | A | -0.010 | **7.71E-08** | 0.006 |
| Substance use | Alcohol intake frequency | 453,069 | 1.05E-14 | rs9814516 | T | -0.026 | **1.09E-12** | 0.011 |
| Substance use | Alcohol usually taken with meals | 235,563 | 1.27E-08 | rs12493621 | C | 0.008 | **3.85E-08** | 0.013 |
| Substance use | Average weekly beer plus cider intake | 324,232 | 6.43E-09 | rs9824301 | C | -0.013 | **8.45E-09** | 0.010 |
| Substance use | Average weekly red wine intake | 323,634 | 5.01E-07 | rs382210 | G | -0.015 | **1.11E-08** | 0.010 |
| Substance use | Current tobacco smoking | 453,155 | 3.76E-11 | rs56262138 | A | -0.005 | **2.60E-08** | 0.007 |
| Substance use | Ever smoked | 451,902 | 1.01E-20 | rs6790699 | A | 0.009 | **6.39E-16** | 0.014 |
| Substance use | Ever taken cannabis | 146,758 | 2.46E-18 | rs62263912 | G | 0.029 | **7.44E-16** | 0.046 |
| Substance use | Frequency of alcohol use, derived for PMID 30874500 | 453,070 | 1.05E-14 | rs9814516 | T | -0.026 | **1.09E-12** | 0.011 |
| Substance use | Frequency of drinking alcohol | 146,785 | 4.44E-07 | rs9832119 | T | -0.025 | **1.06E-06** | 0.016 |
| Substance use | Lifetime cannabis use, derived for PMID 30150663 | 146,758 | 9.11E-16 | rs62263912 | G | 0.011 | **2.63E-12** | 0.033 |
| Substance use | Light smokers, at least 100 smokes in lifetime | 124,240 | 8.13E-10 | rs62253088 | T | 0.011 | **2.49E-07** | 0.020 |
| Substance use | Past tobacco smoking | 417,577 | 4.51E-22 | rs6780346 | C | -0.024 | **7.54E-18** | 0.018 |
| Substance use | Smoking initiation, derived for PMID 30643251 | 301,588 | 2.44E-21 | rs62263910 | G | 0.008 | **3.26E-14** | 0.019 |

\*For binary traits, the effective sample size is given (determined using $N_{\text{eff}} = 4 / \left( \frac{1/N_{\text{cases}}}{1/N_{\text{controls}}} \right)$ ).

**Figure 1.**
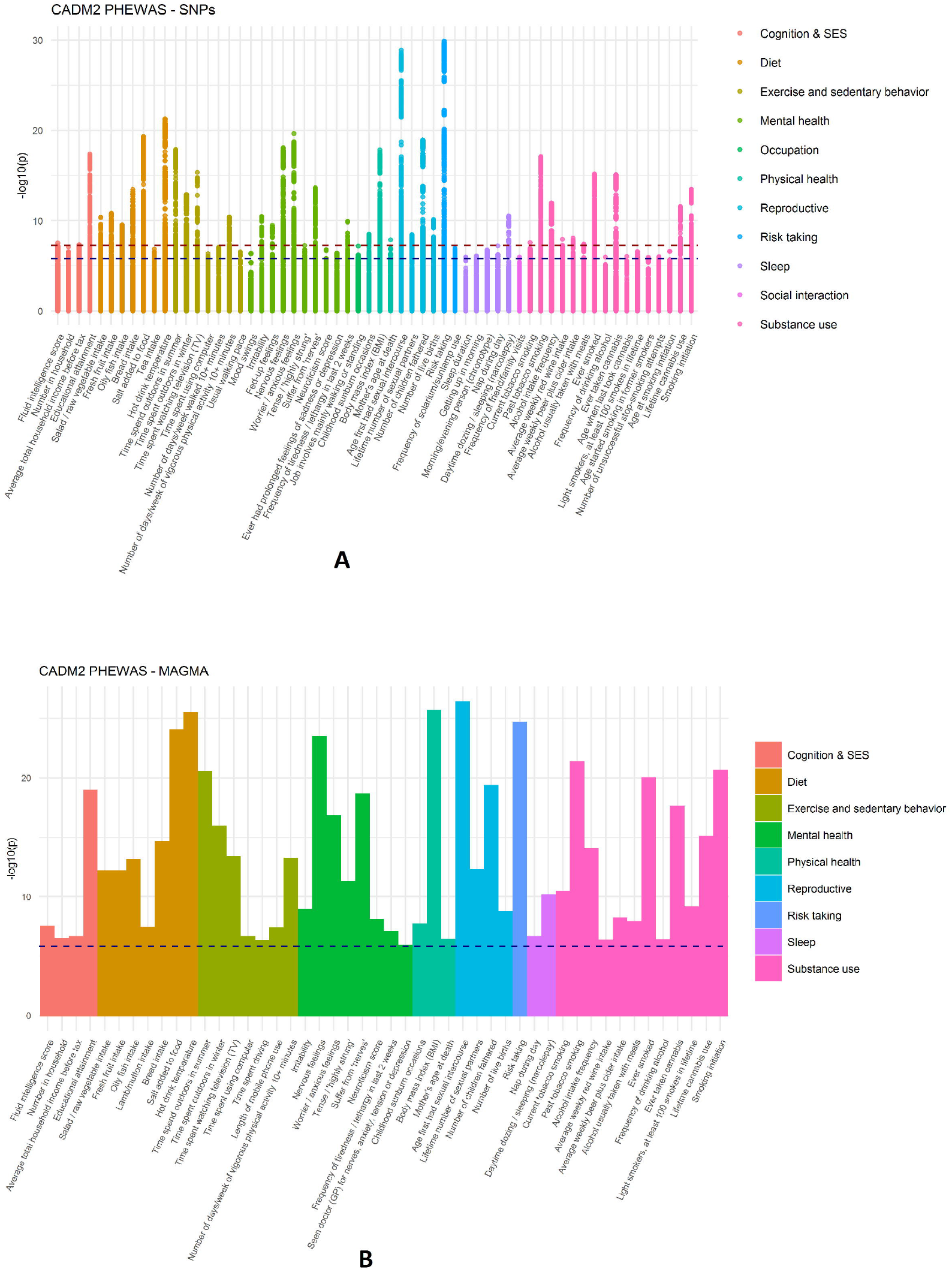
PheWAS results. Panel **a**) shows the subset of significant associations of the SNP-based test (61 out of 242 traits). The x-axis shows the traits (colored by trait category) and the y-axis the p-values of the association. Each dot represents a SNP association. SNPs exceeding the red horizontal line have a *p*-value significant at a genome-wide threshold of *p*=5E-08. The blue horizontal line represents the suggestive threshold of *p*=1.55E-06. Full SNP-based results are given in Supplementary Figure 1. Panel **b**) shows the subset of significant results of the MAGMA gene-based test (51 out of 242 traits), with *p*-values on the y-axis. The red dotted line represents a threshold of *p*=2.62E-06. The full gene-based results are depicted in Supplementary Figure S2.

The SNPs that showed the highest number of significant trait-associations (with a maximum of 30 traits at *p*<1.55E-6, Table S4) clustered around loci at 85.53 and 85.62 Mb. As can be seen in Figure 2, most SNPs that were independently (LD R^2^<0.01, distance >250kb) significantly associated with at least one trait cluster in the middle of the gene, a region rich in eQTLs.

**Figure 2.**
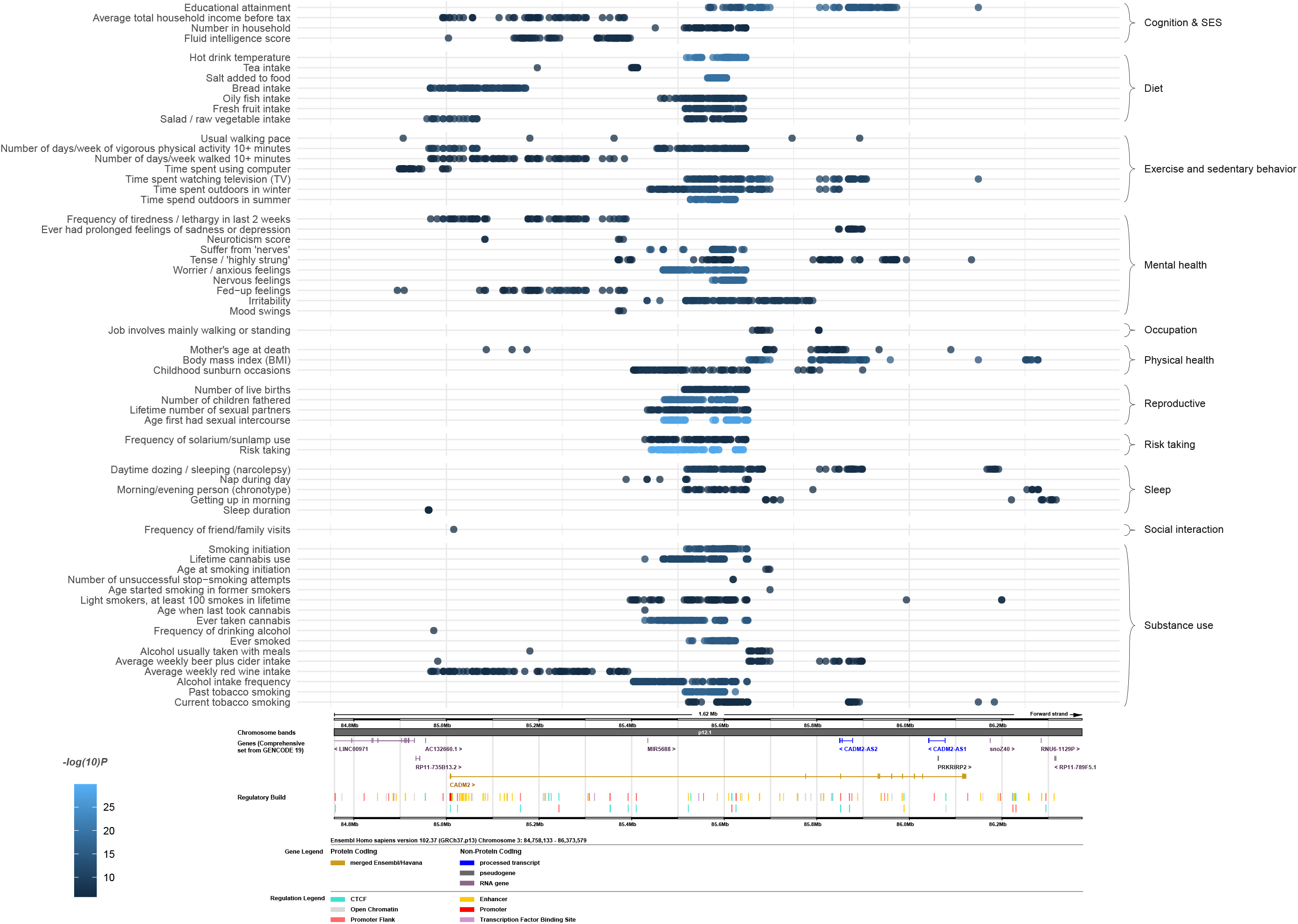
The top 100 most significant SNPs for each trait with at least 1 significant SNP. The x-axis represents the base pair position, and the panel below shows information on the *CADM2* transcripts as derived from https://www.ensembl.org/.

As shown in Figure S3, the high number of associations discovered for *CADM2* was exceptional. Most of the 50 comparison genes contained fewer than 5 trait associations, with an average of 2.8 associated traits per gene and a maximum of 13 (as compared to 38 for *CADM2*; Table S6).

This PheWAS showed that *CADM2* was involved in a wide spectrum of traits, thereby replicating and extending on previous findings. Interestingly, comparison with 50 other genes showed that this number of trait-associations was exceptionally high, emphasizing the distinctive role of *CADM2* in psycho-behavioral traits. Many of the associations we found have been reported in previous literature (Table S4). However, some specific associations, such as those with specific dietary traits, daytime sleeping, number of live births, and mother’s age at death have not been reported before (to our knowledge). Table S5 summarizes putatively novel SNPs for which no phenotypic associations have been reported before, providing an indication of which may be functionally involved in biological pathways. The variance explained by *CADM2* was highest for lifetime cannabis use, followed by number of children fathered, age at first sexual intercourse, and risk taking. Overall, effect sizes were small (less than 0.05% for cannabis initiation), in range with what is normally found for single variants. Few associations were found in the social interaction, sleep, traumatic experiences, and occupational categories. Also, there were not many mental health traits that showed an association (8 out 53 traits). It is interesting to note the significant associations with worry and nervousness-like traits in the absence of association with (other) depression- and anxiety-related traits. There may be something specific to these seemingly overlapping traits, translating to distinct biological pathways.

It needs to be noted that sample sizes for the phenotypes differed substantially (from N=12,211 to 453,349), and as such, it is possible that the pattern of associations was driven in part by differences in power. The correlation between sample size and *p*-value of the gene-based test was moderate and significant, *r*=-.38 (*p*=1.42E-9) showing that well-powered traits were more likely to result in a significant association. It is clear that high power was a requirement: the effect sizes of *CADM2* were diminutive, as is expected for single genes and complex traits. Also, our tests were limited to the psycho-behavioral traits measured in the UK-Biobank; inclusion of more measures, such as longitudinal or non-self-report measures could contribute to a more complete picture. Still, the range of tested traits was quite broad and enabled us to discern interesting patterns.

More research is needed to elucidate these links between *CADM2* and this spectrum of psycho-behavioral traits in terms of neurobiological mechanisms. For example, it could be that *CADM2* is important for the learning aspects of behavior, given its role in synaptic connectivity. Speculatively, *CADM2* could then contribute to reward-learning and associative learning, giving rise to risky behavior and substance use ^18^.

This study presents the first comprehensive and rigorous test of associations between *CADM2* and psycho-behavioral traits, showing strong associations for a wide range of traits (many akin to health behavior). Results could be used as starting point for future research into the function of *CADM2*. Research on the trait-associations and function of *CADM2* will further our understanding of the biology of behavior.

## Data Availability

UK Biobank data are available for researchers upon request at UK Biobank. Summary statistics from the current project are provided in the supplementary materials.

## Acknowledgments

KJHV and AA are supported by the Foundation Volksbond Rotterdam. AA is supported by ZonMw grant 849200011 from The Netherlands Organisation for Health Research and Development. This project was supported by a grant from Amsterdam Neuroscience (2019). We acknowledge SURFsara for the usage of the Cartesius cluster computer (supported by NWO, EINF-457). This project was conducted under UK-Biobank application 40310.

## Author contributions

ZC and JAP were responsible for the analyses, under supervision of AA and KJHV. JAP, ZC, AA, and KJHV wrote the manuscript. The study was conceived by KJHV and AA. Figures were created by AA. All authors read the manuscript draft and provided input.

## Competing interests statement

The authors do not report any competing interests.

## Notes

### Competing Interest Statement

The authors have declared no competing interest.

### Author Declarations

The UK Biobank was approved by the relevant ethics committees https://www.ukbiobank.ac.uk/learn-more-about-uk-biobank/about-us/ethics. This project was evaluated and approved by the Montreal Heart Institute Institutional Review Board under project #2019-2435. All necessary patient/participant consent has been obtained and the appropriate institutional forms have been archived.

