## Supplementary Figures for "The *CADM2* gene and behavior: A phenome-wide scan in UK-Biobank"

**Supplementary Figure S1.** Full results of the SNP-PheWAS. The x-axis shows the traits (colored by trait category)and the y-axis the *p*-values of the association. Each dot represents a SNP association. SNPs exceeding the red horizontal line have a *p*-value significant at a genome-wide threshold of *p*=5E-08. The blue horizontal line represents the suggestive threshold of *p*=1.56E-06.

**Supplementary Figure S2.** Full results for the gene-PheWAS. The x-axis shows the traits (colored by trait category)and the y-axis the *p*-values of the MAGMA association.  Traits exceeding the blue line have a *p*-value significant at a threshold of *p*=2.62E-06 (corrected for the number of genes in the genome).

**Supplementary Figure S3.** Number of significant trait associations per comparison gene (with SNP associations at *p*<5E-08). In blue the *CADM2* gene, the only one with more than 13 trait associations. See Table S6 for the results per comparison gene.

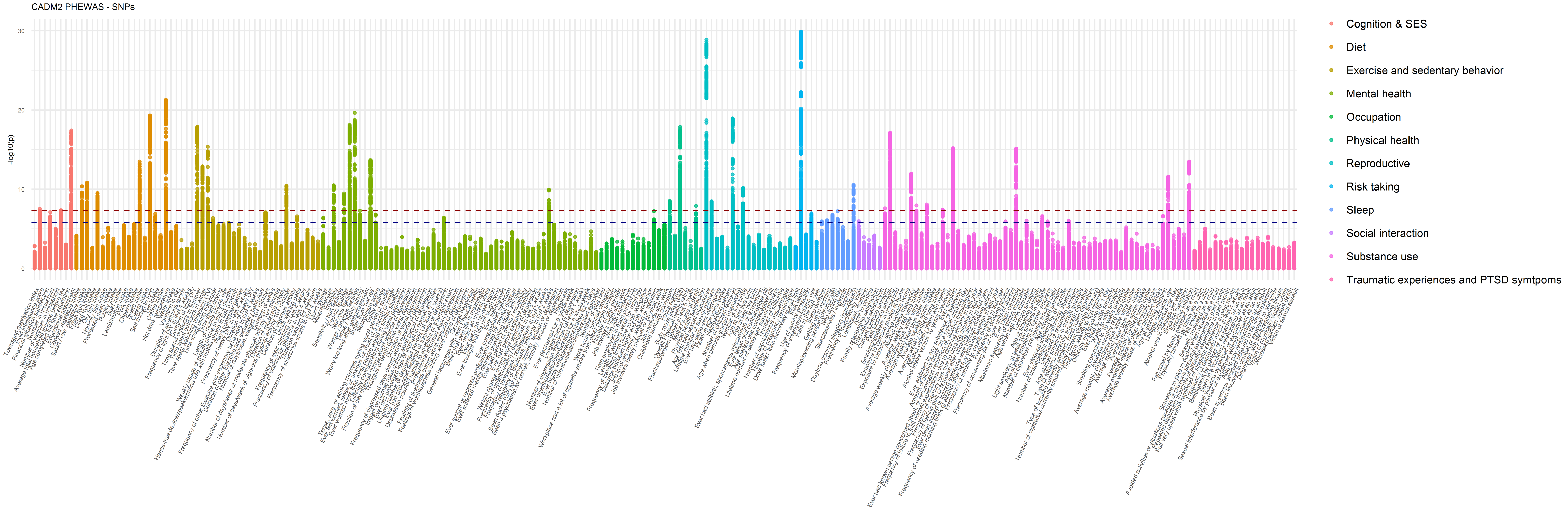

CADM2 PHEWAS - SNPs
-log10(p)
30
20
10
0
Cognition & SES
Diet
Exercise and sedentary behavior
Mental health
Occupation
Physical health
Reproductive
Risk taking
Sleep
Social interaction
Substance use
Traumatic experiences and PTSD symtpoms

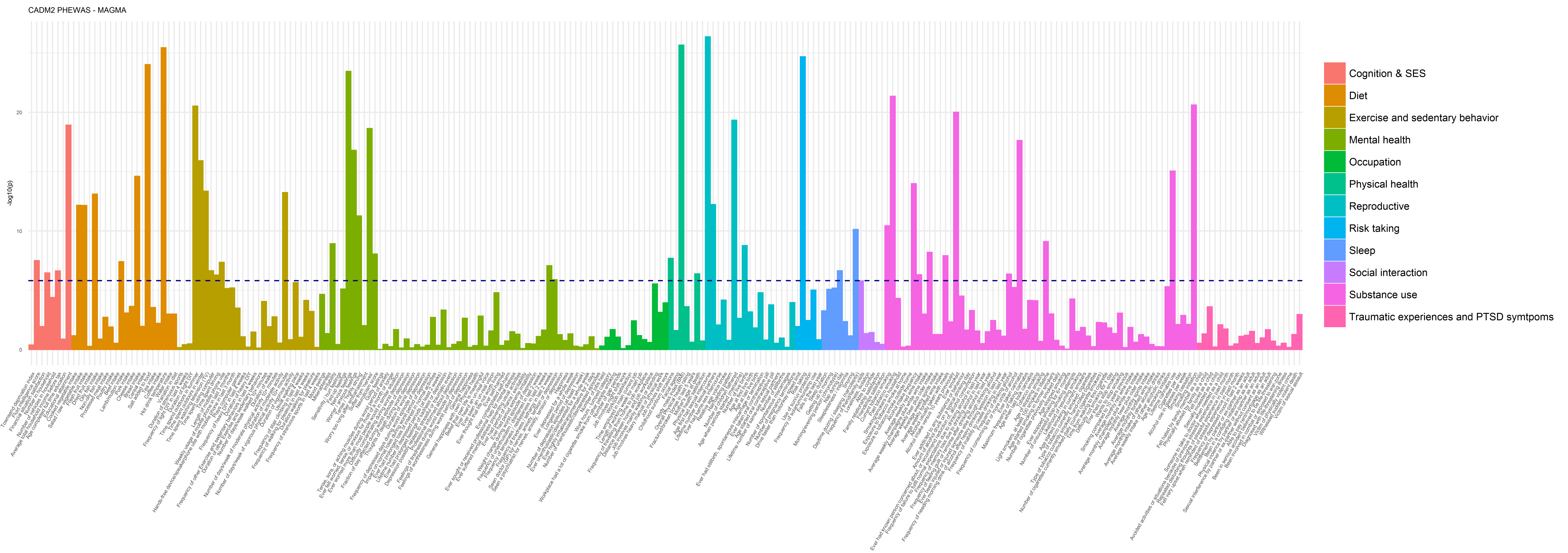

CADM2 PHEWAS - MAGMA
-log10(p)
0
10
20
Cognition & SES
Diet
Exercise and sedentary behavior
Mental health
Occupation
Physical health
Reproductive
Risk taking
Sleep
Social interaction
Substance use
Traumatic experiences and PTSD symtpoms

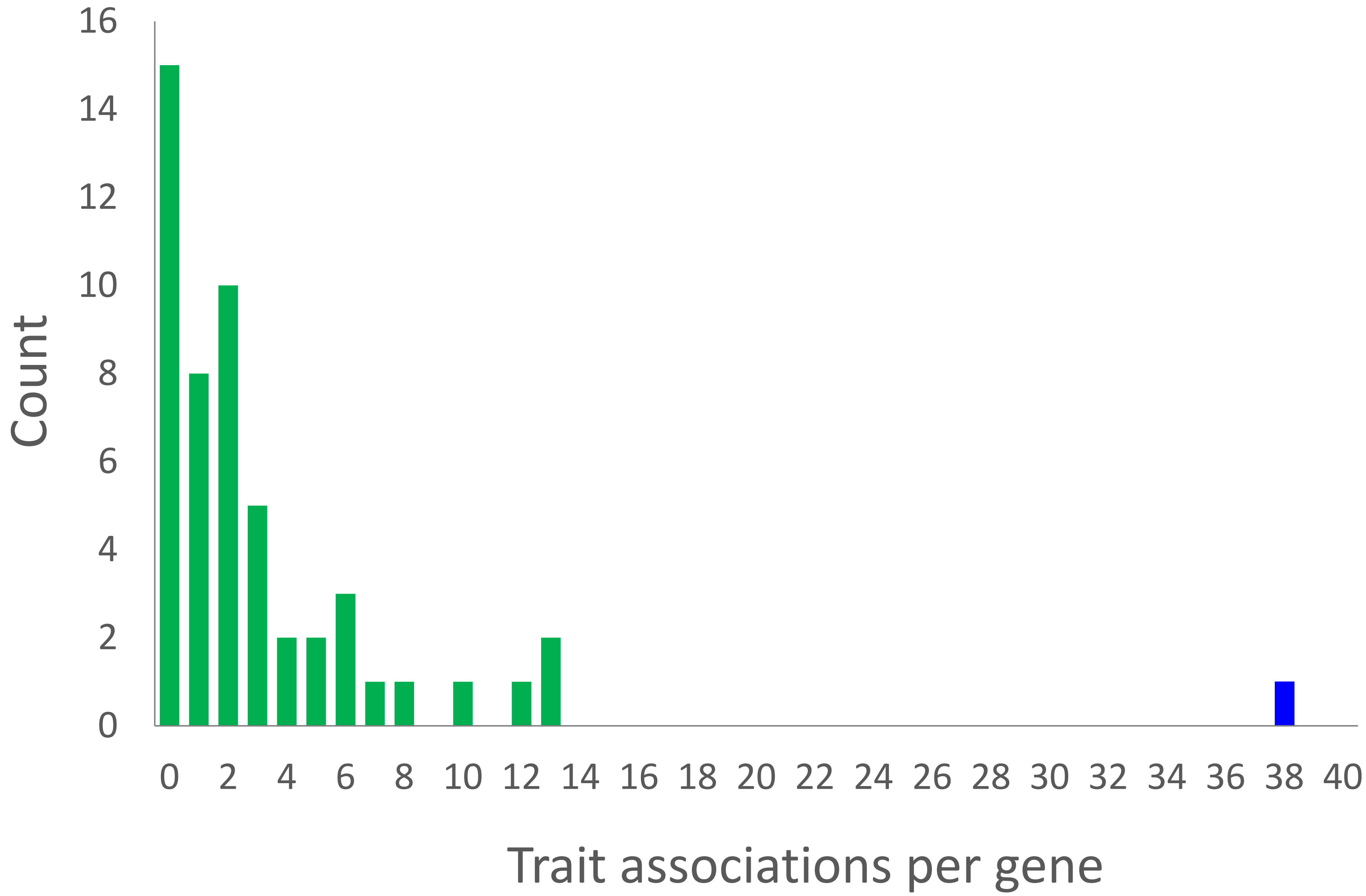
Count
16
14
12
10
8
6
4
2
0
0 2 4 6 8 10 12 14 16 18 20 22 24 26 28 30 32 34 36 38 40
Trait associations per gene
